# Leveraging video data from a digital smartphone autism therapy to train an emotion detection classifier

**DOI:** 10.1101/2021.07.28.21260646

**Authors:** Cathy Hou, Haik Kalantarian, Peter Washington, Kaiti Dunlap, Dennis P. Wall

## Abstract

Autism spectrum disorder (ASD) is a neurodevelopmental disorder affecting one in 40 children in the United States and is associated with impaired social interactions, restricted interests, and repetitive behaviors. Previous studies have demonstrated the promise of applying mobile systems with real-time emotion recognition to autism therapy, but existing platforms have shown limited performance on videos of children with ASD. We propose the development of a new emotion classifier designed specifically for pediatric populations, trained with images crowdsourced from an educational mobile charades-style game: *Guess What?*. We crowdsourced the acquisition of videos of children portraying emotions during remote game sessions of *Guess What?* that yielded 6,344 frames from fifteen subjects. Two raters manually labeled the frames with four of the Ekman universal emotions (happy, scared, angry, sad), a “neutral” class, and “n/a” for frames with an indeterminable label. The data were pre-processed, and a model was trained with a transfer-learning and neural-architecture-search approach using the Google Cloud AutoML Vision API. The resulting classifier was evaluated against existing approaches (Microsoft’s Azure Face API and Amazon Web Service’s Rekognition) using the standard metrics of F1 score. The resulting classifier demonstrated superior performance across all evaluated emotions, supporting our hypothesis that a model trained with a pediatric dataset would outperform existing emotion-recognition approaches for the population of interest. These results suggest a new strategy to develop precision therapy for autism at home by integrating the model trained with a personalized dataset to the mobile game.

## INTRODUCTION

Autism spectrum disorder (ASD) is a neurodevelopmental disorder affecting one in 40 children in the United States and is associated with impaired social interactions, restricted interests, and repetitive behaviors [1-2]. While there is no cure, studies have shown the efficacy of Applied Behavioral Analysis (ABA) therapy, if administered at a young age and customized to address the child’s unique deficits [3-4]. However, caring for a child with ASD can generate a financial burden on the family [5]. Additionally, the increasing prevalence of the condition is resulting in a short supply of certified specialists, further hindering treatment options [6].

We developed *Guess What?* [7-9], a charades-style mobile game that delivers social training to children with ASD at home, to mitigate the high costs and shortage of traditional interventions. To play the game, the child interprets and acts out prompts that are displayed on the screen while the caregiver is tasked with guessing the prompt. Multiple decks and prizes tailor to the child’s preferences and help increase engagement.

*Guess What?* incorporates two teaching methods based on ABA principles: Discrete Trial Training (DTT) and Pivotal Response Treatment (PRT). DTT breaks down the skill into discrete trials that build up the skill step by step [10]. Each trial follows a specific set of steps consisting of an antecedent, prompt, response, reinforcement, and brief pause [10]. PRT is less structured and initiated by the child, emphasizing natural reinforcement and targeting pivotal areas of a child’s development instead of specific behaviors [11]. Multiple studies suggest that DTT helps improve emotion recognition and expression [10] and PRT enhances communication skills in children with ASD [11].

Previous studies have demonstrated the promise of applying mobile systems with real-time emotion recognition to ABA therapy [12-18]. Integrating an automatic emotion classifier into *Guess What?* will provide supplemental reinforcement to the caregiver and allow for the development of additional features integral to ABA therapy: adapting prompts and difficulty to target the child’s specific deficits and offering appropriate visual cues to assist the child [3].

However, existing emotion recognition platforms are not optimized for research on children [19-20] as a result of being trained on datasets in which pediatric populations are highly underrepresented such as the CIFAR-100, ImageNet [21], Cohn-Kanade Database [22] and Belfast-Induced Natural Emotion Databases [23]. *Guess What?* can serve as a data acquisition tool and aggregate emotive videos for autism research that can be used to train a more effective automatic emotion recognition platform. The use of data collected from mobile devices, such as the built-in camera, allow for continuous phenotyping and repeat diagnoses in home settings [24-39]. This motivates the development of a new emotion classifier designed specifically for pediatric populations, trained with images crowdsourced from *Guess What?*.

## METHODS

### Game Design

*Guess What?* is available for both Android and iOS platforms [7-9]. The child begins by selecting one of the following themed decks to play: animals, emoji, faces, gestures, jobs, objects, sports, chores, and a special deck created for toddlers. The caregiver will hold the device outwards with the screen facing the child. During the 90-second game session, the child acts out the prompt displayed on the screen while the caregiver guesses. If the caregiver’s guess is correct, the child prompts the caregiver, who tilts the phone. The game then rewards the child with a point, resulting in another image appearing on the screen. This process repeats for a fixed amount of time. The entire game session is recorded using the front-facing camera on the device, focusing on the child’s actions. If the user grants permission to share this footage, the video is uploaded to a secure and encrypted Amazon Web Services S3 bucket. This data upload and storage process is fully compliant with the Stanford University’s High-Risk Application security standards. Additional metadata included with the video includes the prompts used in the session, timing logs, and the number of points awarded.

### Data Collection

Figure 1 illustrates the data aggregation process. The two decks that are the most closely associated with emotion recognition and expression are the *emoji* and *faces* decks. These decks contain emoticons (cartoon representations of facial emotions) and real images of children expressing various emotions, respectively. Using crowdsourced videos from fifteen subjects remotely playing these two decks subsampled at 5 frames per second (FPS), a dataset consisting of 6,344 frames was built.

**Figure 1.**
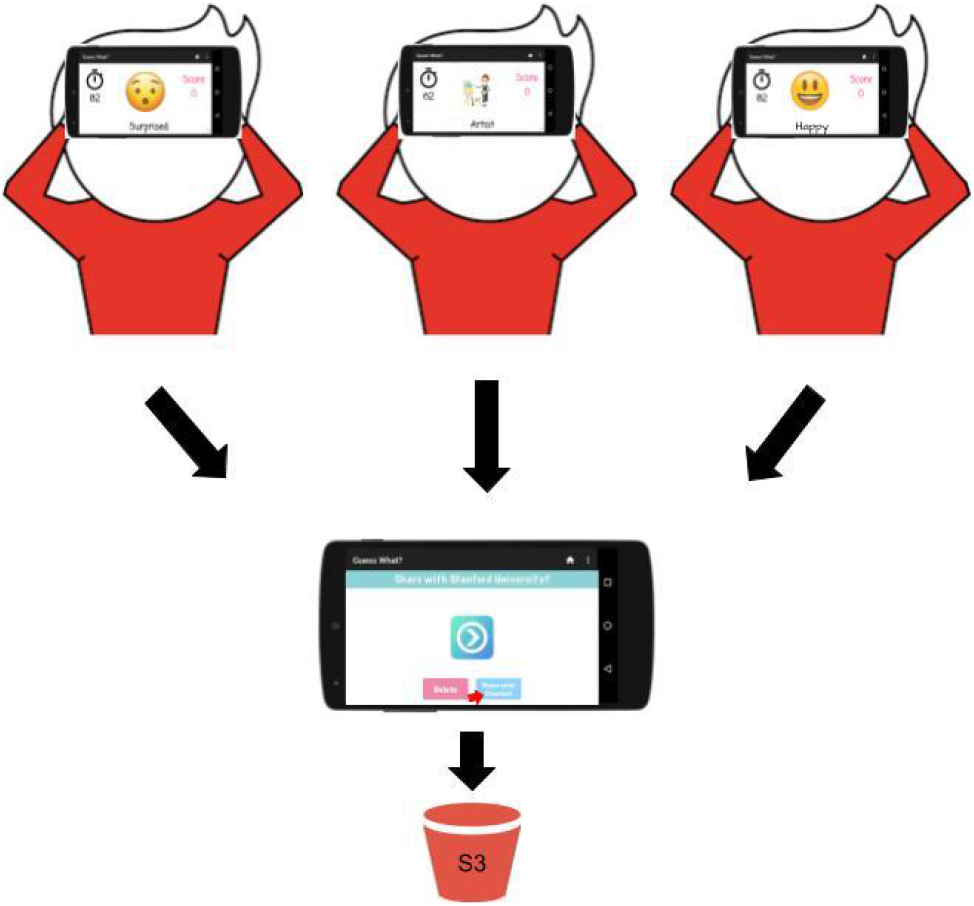
Crowdsourced videos taken during game sessions are stored in an Amazon S3 bucket (with participant’s consent).

### Data Processing

To establish ground truth, two raters manually labeled each of the 6,344 frames with either one of four Ekman universal emotions (happy, sad, scared, angry) [40] or a neutral label. In cases where there were no faces in the frame or the face was too blurry to discern, the raters labeled the frame with “n/a.” To filter the data, all frames with rater disagreement or labeled as “n/a” were discarded. Faces were then extracted from the remaining frames using the OpenCV library [41], yielding 757 frames. Figure 2 is a confusion matrix illustrating the distribution of the raters’ labels. The Cohen’s Kappa statistic for inter-rater reliability [42], a metric which accounts for agreements due to chance, was 0.8, indicating a high level of reliability between the two raters.

**Figure 2.**
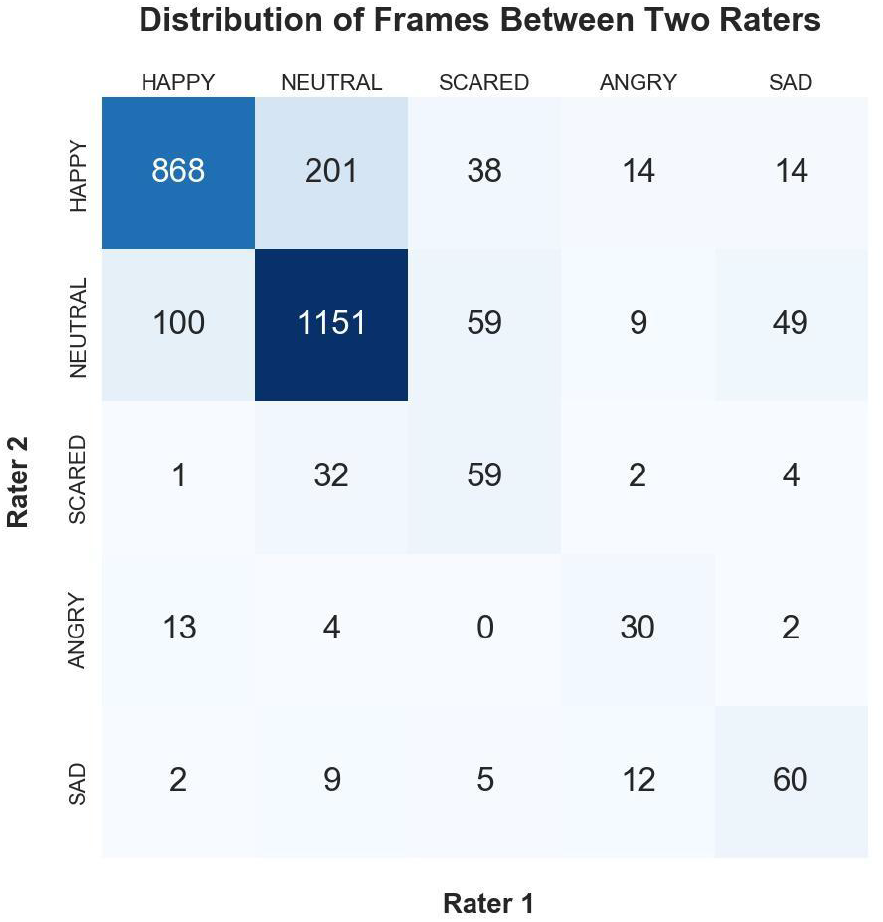
Confusion matrix of the two raters’ emotion labels.

### Classifier Training

The proposed emotion classifier was trained using Google Cloud’s AutoML pipeline, which leverages Google’s transfer learning and neural architecture search technologies to automate the determination of the strongest network architecture and optimal hyperparameter configurations to minimize the loss functions [43-44]. Due to an uneven distribution of emotions, data augmentation methods from the Imgaug library [45] were performed to increase the number of viable frames to 989, with roughly 200 corresponding to each of the five emotions. The specific methods performed included horizontally flipping the image, cropping the image, blurring the image, improving or worsening contrast, adding Gaussian noise to the image, brightening or darkening the image, and applying affine transformations to the image, all performed in random order and of varied magnitudes [45]. 861 frames were used for training and validation, and the remaining 128 frames were used for testing. Figure 3 illustrates the data processing and classifier training procedure.

**Figure 3.**
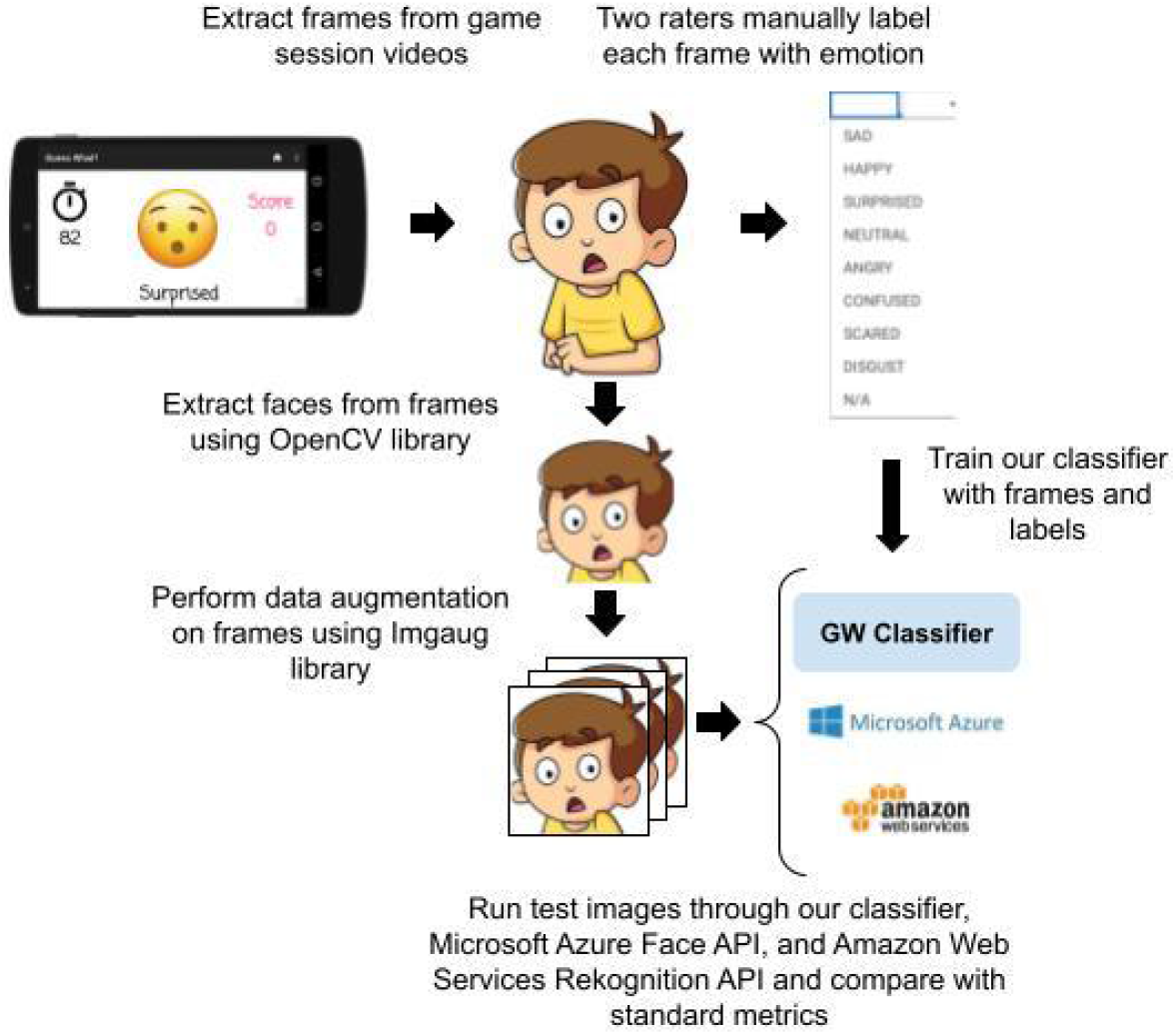
The classifier’s training procedure.

### Data Analysis

To evaluate the performance of the models, the F1 score was calculated as follows:

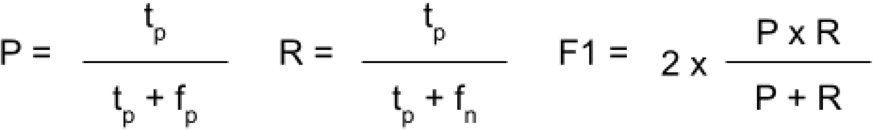

P stands for precision, R stands for recall, F1 stands for F1 score, t_p_ stands for true positive, f_p_ stands for false positive, and f_n_ stands for false negative.

## RESULTS AND DISCUSSION

The proposed classifier was compared to two existing emotion recognition platforms: Microsoft’s Azure Face API [46] and Amazon Web Services’ Rekognition [47]. The same 128 frames that were tested on the proposed classifier were tested on these two classifiers. Figure 4b and 4c show that these classifiers can recognize happy and neutral but perform poorly on angry, sad, and scared classes. These results suggest the need for a new classifier that demonstrates stronger performance across all emotions for pediatric populations.

**Figure 4.**
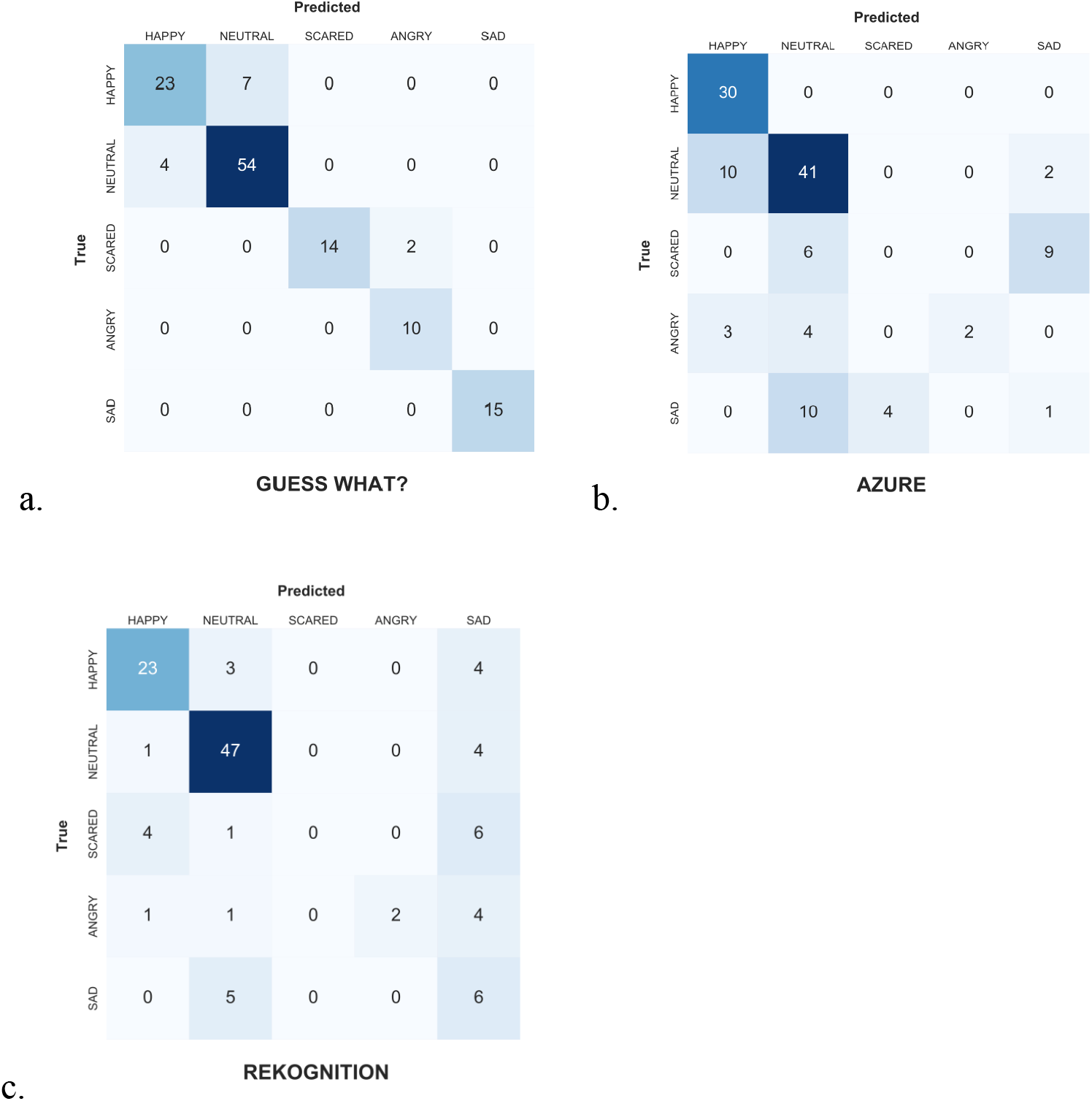
Confusion matrices of the proposed classifier, Azure, and Rekognition.

The performance of the proposed classifier is illustrated in Figure 4a. Figure 4a shows that the most discrepancies occurred between differentiating neutral from happy, which contrasts with the performance of the other classifiers. However, this proposed classifier generally showed a very strong performance for all five emotions, especially when compared to the other two existing classifiers.

Figure 5 shows the F1 scores [48] of each classifier separated by emotion. The proposed classifier displayed the highest F1 scores across all but one emotion in comparison to the existing commercial emotion recognition platforms. The one deviation occurred when the Azure classifier had an F1 score of 0.82 for happy, while the proposed classifier had an F1 score of 0.81. However, the Azure classifier had the lowest F1 scores for all of the other classes.

**Figure 5.**
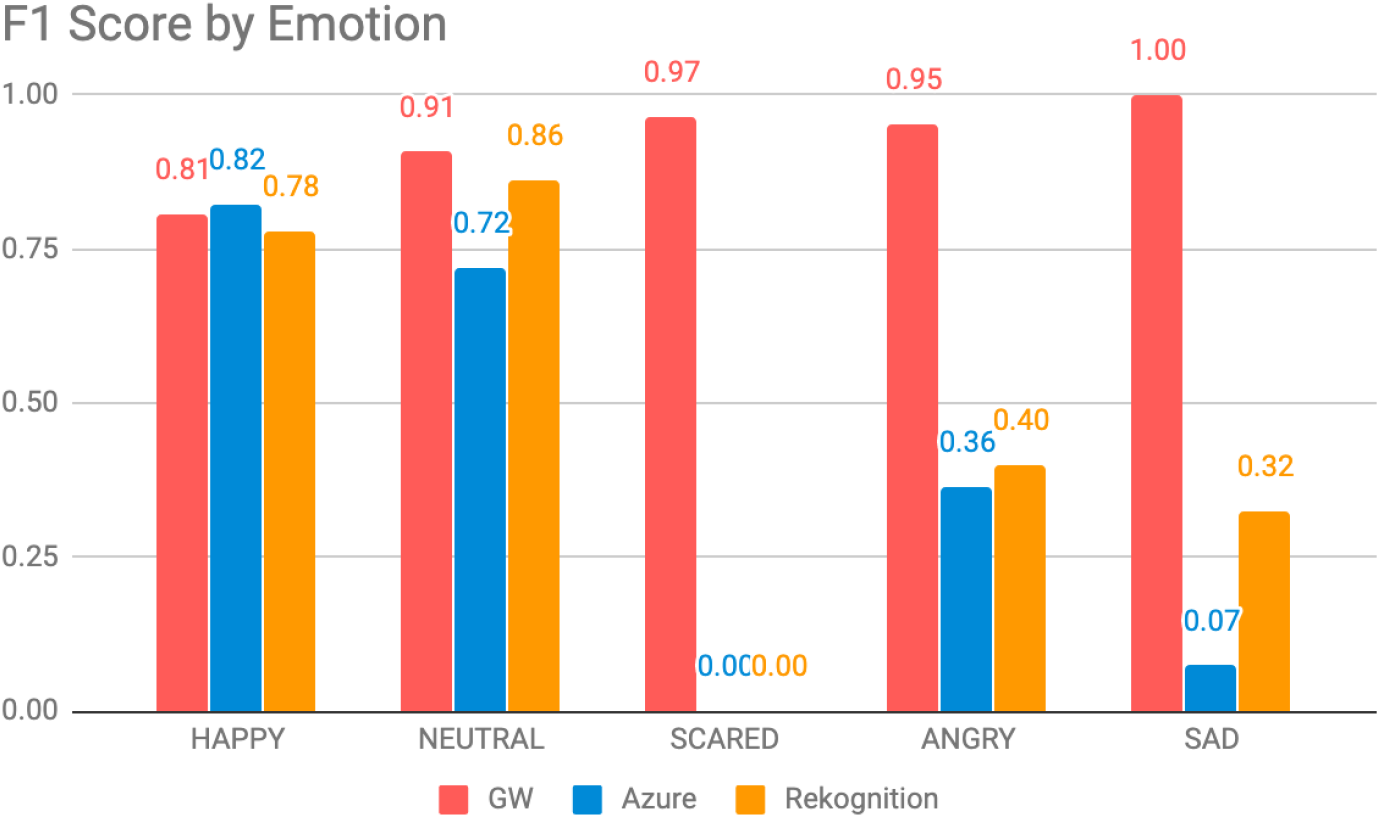
F1 score by emotion by classifier

## CONCLUSION AND FUTURE DIRECTIONS

Both the Azure and Rekognition classifiers performed relatively well on happy and neutral frames but failed with other emotions. In addition, the proposed classifier performed better on scared, angry, and sad frames, while the two existing classifiers demonstrated stronger performance on happy and neutral frames. Because heavy data augmentation procedures had to be performed on the scared, angry, and sad classes to evenly distribute the training set, these results suggest that overfitting may have occurred with the proposed classifier. Generating a more diverse dataset of frames to begin with may alleviate this issue and will be addressed in future work. However, due to the personalized nature of classifiers, perhaps the overfitting is useful in this case. As a result of the limited training set and contrasting performance of the classifiers, in future work, a transfer learning approach will be taken to improve the performance of the proposed classifier across all emotions: the five emotions addressed in this study as well as disgust and surprise which are two other Ekman emotions. Additionally, this emotion classifier generalized to children with ASD will be integrated into *Guess What?* to provide supplemental reinforcement and allow for the development of new features including adapting the game to target specific deficits and providing appropriate guiding feedback.

## Data Availability

A subset of the data that support the findings of this study are available on request from the corresponding author, CH. The data are not publicly available due to their containing information that could compromise the privacy of research participants.

## ACKNOWLEDGEMENTS

These studies were supported by awards to DW by the National Institutes of Health (1R21HD091500-01 and 1R01EB025025-01). Additionally, we acknowledge the support of grants to DW from The Hartwell Foundation, the David and Lucile Packard Foundation Special Projects Grant, Beckman Center for Molecular and Genetic Medicine, Coulter Endowment Translational Research Grant, Berry Fellowship, Spectrum Pilot Program, Stanford’s Precision Health and Integrated Diagnostics Center (PHIND), Wu Tsai Neurosciences Institute Neuroscience: Translate Program, and Stanford’s Institute of Human Centered Artificial Intelligence as well as philanthropic support from Mr. Peter Sullivan. HK would also like to acknowledge support from the Thrasher Research Fund and Stanford NLM Clinical Data Science program (T-15LM007033-35). PW would like to acknowledge support from Mr. Schroeder and the Stanford Interdisciplinary Graduate Fellowship (SIGF) as the Schroeder Family Goldman Sachs Graduate Fellow. Finally, we acknowledge the Stanford Institutes of Medicine Summer Research Program for funding CH.

